# An Exposome-Wide Interaction Study to Identify Hidden Environmental Factors Affecting Susceptible Populations: Application to Chemical-Social Vulnerability Interactions on Cognitive Function

**DOI:** 10.1101/2025.09.17.25335908

**Authors:** Sung Kyun Park, Erika Walker, Laura Arboleda-Merino, Samuel Fansler, Lauren Y. M. Middleton, Xin Wang, Bhramar Mukherjee, Belinda L. Needham, Kelly M. Bakulski

## Abstract

**Background:** Exposome-wide association studies (ExWAS) systematically evaluate numerous environmental exposures, but typically focus on the mean effect, determining whether the association is statistically significant for the average individual. This approach may overlook environmental chemicals that specifically affect susceptible subpopulations. We conducted an exposome-wide interaction study (ExWIS) to identify environmental chemicals associated with cognitive function in the context of social vulnerability.

**Methods:** We examined associations between 147 biomarker-based chemicals and cognitive function among 4,982 adults aged 60 and older in the National Health and Nutrition Examination Survey (NHANES) 1999-2000 and 2011-2014. Social vulnerability index (SVI) was calculated as a composite measure encompassing race/ethnicity, income, education, health insurance, housing type, food security, and employment. Initially, we ran survey-weighted linear regressions between each chemical and cognition, adjusting for SVI, age, sex, smoking status, serum cotinine, fish consumption, NHANES cycle, and urinary creatinine (marginal association models). Subsequently, we repeated these models with a linear interaction term between each chemical and SVI (interaction models).

**Results:** In marginal association models, 16 chemicals including smoking-related compounds, polyaromatic hydrocarbons, and metals were associated with cognitive function (unadjusted *p*<0.05). In interaction models, four additional chemicals–urinary mercury, diethylthiophosphate, perfluorononanoate, and perfluoroundecanoate–not identified in the marginal association models showed significant interactions with SVI.

**Discussion:** This study demonstrates the utility of the ExWIS framework to identify environmental factors that affect susceptible populations but may be missed by the mean effect-based ExWAS approach. Considering interaction effects is essential for accurate environmental risk assessment and identification of neurotoxicants in at-risk populations.

**HIGHLIGHTS:**

- In a nationally representative sample of older adults, chemical exposures and social factors are cross-sectionally associated with cognitive function.
- Proof of concept exposome-wide interaction study (ExWIS) demonstrates jointly incorporating chemicals and social factors identifies exposures relevant for susceptible populations.
- The hypothesis generating ExWIS approach presented here may be broadly relevant for other outcomes and studies.

## 1. INTRODUCTION

Omics data, such as from the genome, methylome, metabolome, and exposome, is increasingly used to identify risk factors and elucidate the biological mechanisms underlying human diseases. Such data is large-scale in terms of the number of exposures (*p*) ranging from hundreds (e.g., targeted chemicals) to hundreds of thousands (e.g., methylome and metabolome) to millions (e.g., genome-wide variants). Unlike hypothesis-driven approaches, omics analyses are data-driven and agnostic, aiming to discover ‘unknown’ potential risk factors. Omics data are typically evaluated systematically using regression models, where each genomic variant or exposure is assessed individually while statistical inference is made with accounting for multiple comparisons, commonly via false discovery rate (FDR) corrections. This approach has been widely used in recent environmental health studies, named as environment-wide association studies or exposome-wide association studies (ExWAS), and which have identified potential risk factors for various health outcomes (Fang et al., 2025; Khodasevich et al., 2025; Lee et al., 2025; Middleton et al., 2025, 2024; Patel et al., 2010; Vrijheid et al., 2020).

One of the major limitations of ExWAS and other omics-wide association studies is their focus on mean effects; in other words, they evaluate whether the association is statistically significant for the average individual in the population. Such analyses may overlook risk in susceptible subpopulations. For instance, some environmental chemicals may have disproportionately adverse effects on certain groups, such as children, older adults, pregnant or postmenopausal women, racial and ethnic minorities, and individuals with low socioeconomic status (Varshavsky et al., 2023). Previous studies have shown that the short-term effects of particulate matter on mortality risk are stronger in older adults and individuals with lower education or income (Bell et al., 2013). Cardiovascular effects of endocrine disrupting chemicals may be greater in men and postmenopausal women, potentially due to the protective effects of estrogen in premenopausal women (Assenza et al., 2025).

Social vulnerability refers to the degree to which individuals, communities, or societies are susceptible to harm from various stressors, such as natural disasters, economic crises, and public health emergencies (Cutter et al., 2003; Mah et al., 2023). This concept has gained importance in public health research, where it is used to identify populations likely to experience negative health outcomes due to exposure to social and economic risk factors (Mah et al., 2023). Social vulnerability is often quantified using the Social Vulnerability Index (SVI), a tool developed by the Centers for Disease Control and Prevention (CDC)/Agency for Toxic Substances and Disease Registry (ATSDR) that assesses factors like socioeconomic status, household characteristics, minority status, housing type, and transportation conditions at the census tract level (CDC/ATSDR, 2024; Flanagan et al., 2018). Populations with higher SVI scores tend to face greater barriers to health, such as limited access to healthcare, poor housing conditions, and poverty, which collectively increase the likelihood of disease and worse health outcomes (Andrew et al., 2008; Wallace et al., 2015). Moreover, these communities are more likely to suffer from environmental injustice, as they often reside in areas with high pollution or industrial activity, which further compounds their health risks (Molchan et al., 2024). The impacts of social vulnerability and chemical exposures on health are often examined in separate studies, though their impacts may be intertwined.

The present study aims to conduct an exposome-wide interaction study (ExWIS), a novel analytic approach that systematically evaluates the associations between many individual environmental chemicals and cognitive function, in the context of social vulnerability. We treat SVI as a potential effect modifier, acknowledging evidence that social stressors can increase susceptibility to health effects of chemical exposures (Gee and Payne-Sturges, 2004; Payne-Sturges et al., 2021). Given the agnostic, data-driven nature of our approach, we did not specify *a priori* hypotheses regarding the direction of chemical-SVI interactions. Identifying chemical exposures relevant in social contexts can help guide policy and practices to support the entire population, including susceptible groups.

## 2. METHODS

### 2.1. Study Population

The US National Health and Nutrition Examination Survey (NHANES) is an ongoing series of cross-sectional surveys managed by the National Center for Health Statistics (NCHS) (CDC/NCHS, 2024a). Each annual survey is an independent, representative sample of the non-institutionalized civilian population in the US. Since 1999, public-use datasets have been published in two-year cycles (e.g., 1999-2000, 2001-2002, etc.). NHANES administered cognitive testing in four cycles: 1999-2000, 2001-2002, 2011-2012, and 2013-2014. We included three of these cycles in this analysis (1999-2000, 2011-2012, and 2013-2014). The 2001-2002 cycle was excluded due to a lack of data for fish consumption, a confounder of interest. All NHANES participants provided informed consent. This secondary data analysis is approved by the University of Michigan Institutional Review Board (HUM00194918). All original datasets used in this analysis are publicly available through the NCHS. Additionally, preprocessed datasets are publicly available through online repositories (Nguyen et al., 2023).

### 2.2. Chemical Biomarkers

A subset of participants in each NHANES cycle provided blood and urine samples during Mobile Examination Center visits. Environmental chemical biomarker levels were measured in these samples by trained laboratory technicians at the National Center for Environmental Health. Laboratory methods and quality control/quality assurance protocols for each measurement and cycle year are publicly available (CDC/NCHS, 2024b). A total of 395 biomarkers were measured in at least one of the three included NHANES cycles. Because these biomarkers were not measured uniformly in all cycles, the samples sizes vary. Data cleaning and exclusion criteria for exposure variables followed previously described methods (Middleton et al., 2025). Chemicals with >50% of measures above the lower limit of detection were included. Within each chemical, measures were first log_2_ transformed, then standardized to a mean of 0 and standard deviation of 1 (Middleton et al., 2025).

### 2.3. Cognitive Function

To measure cognitive function, NHANES administered the Digit Symbol Substitution Test (DSST) to adults aged 60 years and older. In the DSST, the numbers 1 through 9 are paired with symbols such as arrows and squares (Jaeger, 2018). Participants must copy as many symbols as possible in two minutes that correspond to the numbers listed in rows on the test. Each correct symbol was scored as one point for a maximum of 133 possible in NHANES (CDC/NCHS, 2005). The DSST was treated as a continuous variable in the primary analyses. In sensitivity analyses, it was dichotomized at the survey-weighted 25th percentile, indicating mild cognitive impairment (MCI) below this cutoff and normal cognition above (Kim et al., 2015; Middleton et al., 2025).

### 2.4. Social Vulnerability Index (SVI)

Social vulnerability was assessed at the individual level using a modified SVI, adapted from key components of the CDC/ATSDR’s SVI (CDC/ATSDR, 2024; Flanagan et al., 2018) that were available in NHANES questionnaires. The modified index included income, education, health insurance status, race/ethnicity, housing tenure, employment, and food security. Each component was scored separately then summed to create an overall score ranging from 0 (least vulnerable) to 7 (most vulnerable) **(Table 1)**. For example, participants received a score of 0 for income if their household income was above 150% of the poverty line or a 1 if it was at or below 150%. Similarly, for education, participants were scored 0 if they had at least a high school diploma, or 1 if they did not complete high school. Health insurance was categorized as 0 for private insurance, 0.5 for government insurance (Medicaid, Medicare, etc.), and 1 for no insurance. Race/ethnicity was scored 0 if participants identified as non-Hispanic White (NHW) and 1 otherwise. Housing tenure was scored 0 for homeowners and 1 for renters or others. Employment status was scored 0 for those employed and 1 for those unemployed or retired. Finally, food security was scored 0 for full food security and 1 for less than full food security. This modified SVI provided a comprehensive measure of individual level social vulnerability, enabling the study to capture the varying degrees of vulnerability among participants. We treated SVI as a continuous measure in our analyses. For visualization, we categorized SVI into tertiles.

**Table 1.** Social Vulnerability Index (SVI) score components and calculation.

| **Factor** | **Categorization** |
| --- | --- |
| Income | 0 = above 150% of poverty line<br>1 = at or below 150% of poverty line |
| Education | 0 = high school level or above<br>1 = did not complete high school |
| Health insurance | 0 = private insurance<br>0.5 = government insurance (Medicaid, Medicare, other)<br>1 = no insurance |
| Race/ethnicity | 0 = non-Hispanic white (NHW)<br>1 = not NHW |
| Housing tenure | 0 = homeowner<br>1 = renter or other |
| Employment | 0 = employed<br>1 = unemployed or retired |
| Food security | 0 = full food security<br>1 = less than full food security |
| Total SVI <sup>a</sup> | Income + Education + Health Insurance + Race/ethnicity<br>+ Housing tenure + Employment + Food Security |

<sup>a</sup> SVI range: 0-7. Higher score indicates higher vulnerability

### 2.5. Covariate measures

NHANES collected demographic characteristics, health behavior information, and physical measures through interviews and a mobile physical exam center. We identified a set of potential confounders based on prior exposome-wide association studies and literature on cognitive function (Middleton et al., 2025; Park et al., 2021). Age, sex, tobacco use, fish and shellfish consumption, and the cycle year of participation were assessed in interviews. Age in years was top-coded at 85 (prior to 2007) or 80 (2007 onwards) by NHANES to prevent disclosure. Sex was categorized as male or female. Cigarette smoking status was categorized as never (reported smoking fewer than 100 cigarettes), former (reported smoking more than 100 cigarettes without current use), or current (reported current use). The number of various fish and shellfish eaten in the past 30 days, collected in a dietary intake interview, were summed together and categorized into three groups: 0 (no fish consumption reported), 1-3 times, or 4+ times.

Concentrations of serum cotinine (ng/mL), serum creatinine (mg/dL), and urinary creatinine (mg/dL) were measured from samples provided by participants. Serum creatinine was used to calculate estimated glomerular filtration rates (eGFR) based on the Chronic Kidney Disease Epidemiology Collaboration equation (Levey et al., 2009). We did not adjust eGFR values based on race. Kidney function was categorized as normal (eGFR ≥60 ml/min/1.73 m^2^) or abnormal (eGFR <60 ml/min/1.73 m^2^). Serum cotinine and urinary creatinine were log_2_-transformed and z-score standardized. Serum cotinine was included in models for all exposures except smoking-related chemicals, and urinary creatinine was included in all models for urinary biomarkers to control for urine dilution.

### 2.6. Statistical Analysis

Data management and statistical analyses were conducted in R version 4.4.0 (R Core Team, 2024). Analyses were survey-weighted using the package ‘survey’ to incorporate NHANES’ complex sampling methodology and produce nationally representative estimates (Lumley et al., 2024). Code to reproduce this analysis is available on GitHub (https://github.com/bakulskilab/Cognition_SVI_ExWIS).

Participants were included in the analysis if they were at least 60 years old (eligible for the DSST) and had at least one chemical biomarker measurement. We used flow charts to depict the inclusion and exclusion criteria for participants and chemical biomarkers. We compared the distributions of covariates between the included and excluded participants. Descriptive statistics for DSST scores, SVI components, and covariates were calculated as mean and standard deviation for continuous variables and percentages for categorical variables. We compared the distributions of covariates between the three NHANES waves and by cognitive status. We additionally calculated descriptive statistics for the included chemical biomarkers, including arithmetic and geometric means, standard deviations, and percentiles. We calculated Spearman correlation coefficients between pairs of chemical biomarkers and SVI and plotted them in a heatmap matrix.

We imputed missing data for individual SVI components, covariates, and DSST scores using multiple imputation by chained equations (MICE) (van Buuren, 2018; van Buuren and Groothuis-Oudshoorn, 2011). The imputed variables are described in **Supplemental Table 1**. We generated five complete datasets from the MICE procedure. Following guidelines for multiple imputation analysis, all models described below were run with each imputed dataset (producing five separate sets of coefficient estimates) and pooled together using Rubin’s rules to create one set of estimates and measures of significance to report per model (van Buuren, 2018).

To evaluate marginal associations between chemical exposure biomarkers and cognitive function, we first ran separate survey-weighted linear regression models for each individual exposure biomarker. Models were adjusted for age, sex, SVI, smoking status, serum cotinine, urinary creatinine (urinary biomarker models only), fish consumption, and NHANES cycle year. Second, to evaluate SVI as an effect modifier, we added an interaction term between SVI and the chemical exposure measure to all models. We used a conventional significance threshold of p-value of 0.05 as well as Benjamini-Hochberg false discovery rate (FDR)-corrected p-values to account for multiple comparisons (Benjamini and Hochberg, 1995). We visualized results using a set of three volcano plots to depict the chemical coefficient in the marginal association model, the chemical coefficient in the interaction model, and the SVI-chemical interaction coefficient in the interaction model. For any models with significant SVI-chemical interactions, we also created interaction plots to visualize associations between the chemical exposures and DSST stratified by levels of SVI. These plots were made with complete-case data, rather than the MICE data.

### 2.7. Sensitivity Analyses

We conducted three sensitivity analyses to further investigate these associations. First, we restricted the sample to participants with normal kidney function based on eGFR measures to account for the potential influence of diminished kidney function on chemical biomarker concentrations. Second, we repeated the analysis using binary cognitive status (MCI vs. no impairment) as the outcome in modified Poisson regression models. Last, we ran a two degrees of freedom (2-df) Wald test to compare models with and without the chemical exposure and chemical exposure-SVI interaction terms to replicate an approach commonly used in gene-environment interaction studies (Boonstra et al., 2016; Kraft et al., 2007).

## 3. RESULTS

### 3.1. Population Characteristics

There were 4,982 participants and 147 chemical biomarkers included in this analysis (**Supplemental Figure 1**). Included participants (unweighted N=4,982) had an average age of 69.8 years (SD=7.0) (**Table 2**). The mean DSST score was 50.4 (SD=17.4) and the mean SVI was 1.9 (SD=1.5). Those excluded due to missing chemical measurements were older and had a lower DSST score and higher SVI (**Supplemental Table 2**). Participants with MCI (N=1,493) tended to be older and have higher SVI, higher serum cotinine levels, and lower fish consumption compared to those with no cognitive impairment (N=2,768) (**Supplemental Table 3**). Descriptive statistics for chemicals are presented in **Supplemental Table 4**. **Figure 1** shows a correlation heatmap of chemical exposure biomarkers and SVI. In general, high correlations were observed among chemicals within the same chemical family, particularly polyaromatic hydrocarbons, polychlorinated biphenyls, and smoking-related compounds. SVI components had weak correlations with chemicals.

**Figure 1.**
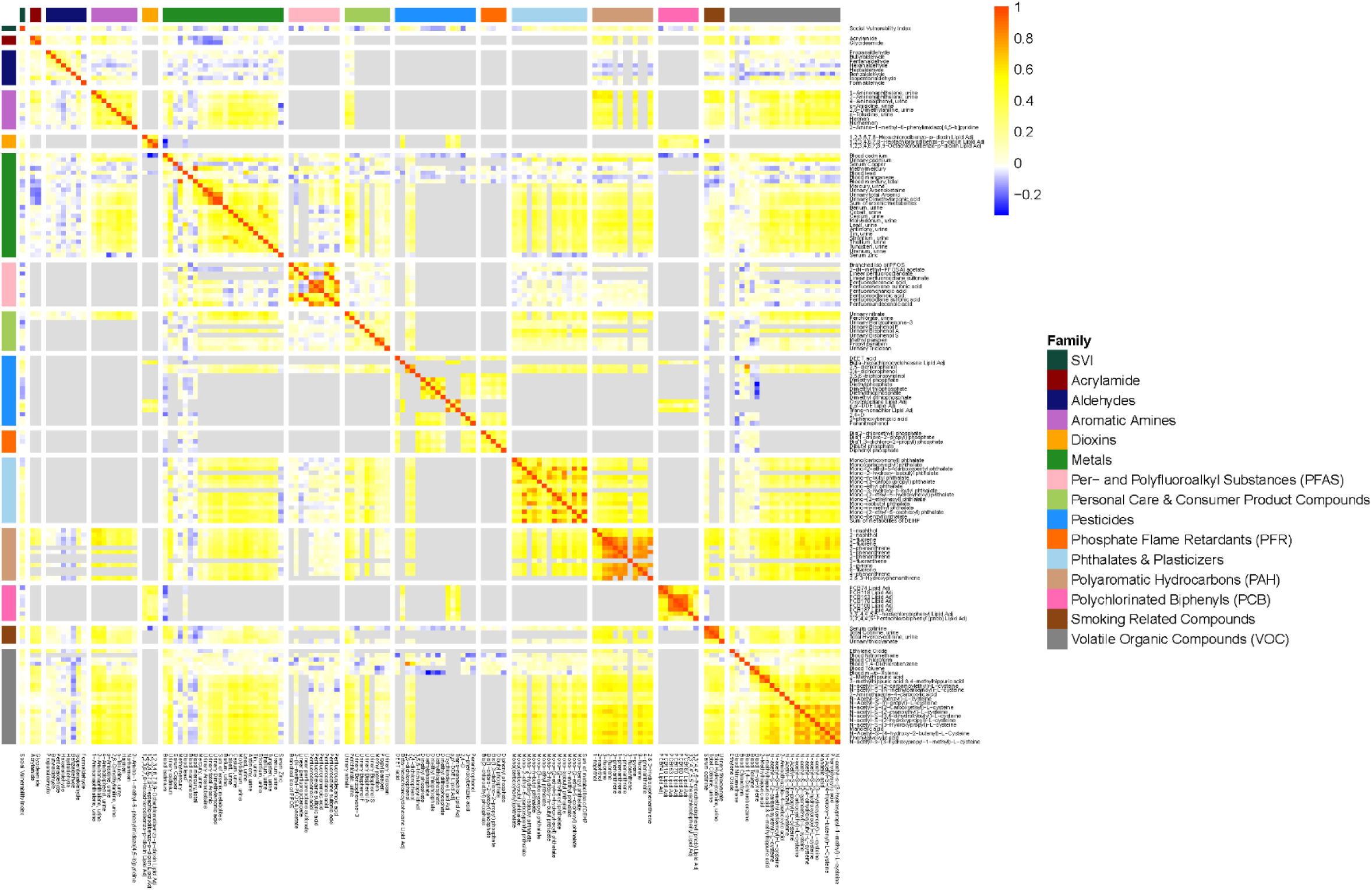
Heatmap of Spearman correlation coefficients between the social vulnerability index (SVI) and 147 included chemical exposures. The variables are grouped by family and color-coded across the outer row and column. The cell color indicates the correlation, with negative correlations in blue shifting to positive correlations in red. A gray cell means that no overlapping participants had those two measures, so no correlation could be calculated.

**Table 2.** Weighted descriptive statistics of participants in the National Health and Nutrition Examination Survey (NHANES) included in the analytic sample, stratified by cycle of participation (N = 4,982).

| **Variable** | **Overall**<br>**(N = 4,982)<sup>1</sup>** | **1999-2000**<br>**(N = 1,562)<sup>1</sup>** | **2011-2014**<br>**(N = 3,420)<sup>1</sup>** | **p-value<sup>2</sup>** |
| --- | --- | --- | --- | --- |
| **Digit Symbol Substitution Test (DSST)** | 50.4 (17.4) | 45.9 (17.9) | 51.9 (16.9) | <0.001 |
| Missing | 721 | 291 | 430 |  |
| **Age (years)** | 69.8 (7.0) | 70.3 (7.4) | 69.6 (6.8) | 0.1 |
| **Sex** |  |  |  | 0.6 |
| Male | 45.0% | 44.5% | 45.2% |  |
| Female | 55.0% | 55.5% | 54.8% |  |
| **Social Vulnerability Index (SVI)** | 1.9 (1.5) | 2.1 (1.6) | 1.9 (1.5) | 0.04 |
| Missing | 621 | 295 | 326 |  |
| **Poverty-income ratio (PIR)** |  |  |  | 0.03 |
| PIR > 1.5 | 74.0% | 66.4% | 76.3% |  |
| PIR ≤ 1.5 | 26.0% | 33.6% | 23.7% |  |
| Missing | 583 | 273 | 310 |  |
| **Education** |  |  |  | <0.001 |
| Completed high school or above | 77.0% | 65.0% | 81.1% |  |
| Did not complete high school | 23.0% | 35.0% | 18.9% |  |
| Missing | 15 | 9 | 6 |  |
| **Race/ethnicity** |  |  |  | 0.5 |
| Non-Hispanic White | 78.2% | 80.3% | 77.5% |  |
| Not Non-Hispanic White | 21.8% | 19.7% | 22.5% |  |
| **Health insurance** |  |  |  | 0.5 |
| Private insurance | 61.7% | 59.8% | 62.4% |  |
| Government insurance | 33.0% | 35.5% | 32.1% |  |
| No insurance | 5.3% | 4.7% | 5.5% |  |
| Missing | 46 | 34 | 12 |  |
| **Housing tenure** |  |  |  | 0.3 |
| Homeowner | 81.7% | 79.5% | 82.5% |  |
| Renter or other | 18.3% | 20.5% | 17.5% |  |
| Missing | 52 | 27 | 25 |  |
| **Food security** |  |  |  | <0.001 |
| Full security | 88.2% | 92.4% | 86.7% |  |
| Less than full security | 11.8% | 7.6% | 13.3% |  |
| Missing | 57 | 33 | 24 |  |
| **Employment** |  |  |  | 0.03 |
| Employed | 27.3% | 23.0% | 28.7% |  |
| Unemployed or retired | 72.7% | 77.0% | 71.3% |  |
| Missing | 2 | 0 | 2 |  |
| **Serum cotinine (ng/mL)** | 35.9 (105.0) | 36.1 (99.6) | 35.9 (106.7) | >0.9 |
| Missing | 295 | 119 | 176 |  |
| **Smoking status** |  |  |  | 0.3 |
| Never smoker | 49.1% | 47.1% | 49.7% |  |
| Former smoker | 39.2% | 39.6% | 39.0% |  |
| Current smoker | 11.8% | 13.3% | 11.3% |  |
| Missing | 7 | 4 | 3 |  |
| **Urinary creatinine (mg/dL)** | 104.8 (67.9) | 107.5 (71.2) | 103.9 (66.7) | 0.4 |
| Missing | 139 | 44 | 95 |  |

| Variable | Overall<br>(N = 4,982)<sup>1</sup> | 1999-2000<br>(N = 1,562)<sup>1</sup> | 2011-2014<br>(N = 3,420)<sup>1</sup> | p-value<sup>2</sup> |
| --- | --- | --- | --- | --- |
| **Fish and seafood consumption in**<br>**past 30 days** |  |  |  | 0.1 |
| 0 times | 14.4% | 15.6% | 13.9% |  |
| 1-3 times | 33.7% | 36.6% | 32.6% |  |
| 4+ times | 51.9% | 47.8% | 53.5% |  |
| Missing | 444 | 53 | 391 |  |
| **Estimated glomerular filtration rate**<br>**(eGFR; ml/min/1.73 m<sup>2</sup>)** |  |  |  | <0.001 |
| eGFR < 60 | 21.3% | 12.0% | 24.5% |  |
| eGFR ≥ 60 | 78.7% | 88.0% | 75.5% |  |
| Missing | 288 | 92 | 196 |  |

<sup>1</sup>Mean (SD); %; unweighted N missing
<sup>2</sup>t-test adapted to complex survey samples; chi-squared test with Rao & Scott's second-order correction

### 3.2. Marginal Association Models

In regression models without inclusion of the interaction term, 16 biomarkers were associated with cognitive function at unadjusted p<0.05, including smoking-related compounds, polyaromatic hydrocarbons, volatile organic compounds, metals, organophosphate flame retardants, pesticides, and per- and polyfluoroalkyl substances (PFAS) (**Figure 2, Panel A; Table 3**). For example, a doubling of serum cotinine concentration was associated with ‒1.93 points difference in DSST score (95% CI ‒3.06, ‒0.80). Benzophenone-3 (urine) was associated with 1.71 points higher DSST (95% CI 0.61, 2.81) per doubling. Additional chemical biomarkers with unadjusted p<0.01 include total cotinine (urine) (β=‒3.52 (95% CI ‒4.89, ‒ 2.15), p=0.005), total hydroxycotinine (urine) (β=‒3.60 (95% CI ‒5.08, ‒2.12), p=0.007), and N-acetyl-s-(3-hydroxypropyl-1-methyl)-L-cysteine (urine) (β=1.64 (95% CI 0.52, 2.76), p=0.0099). However, none of these associations reached statistical significance at FDR-corrected p-values <0.05. Full results for all exposures, regardless of statistical significance, are in **Supplemental Table 5**.

**Figure 2.**
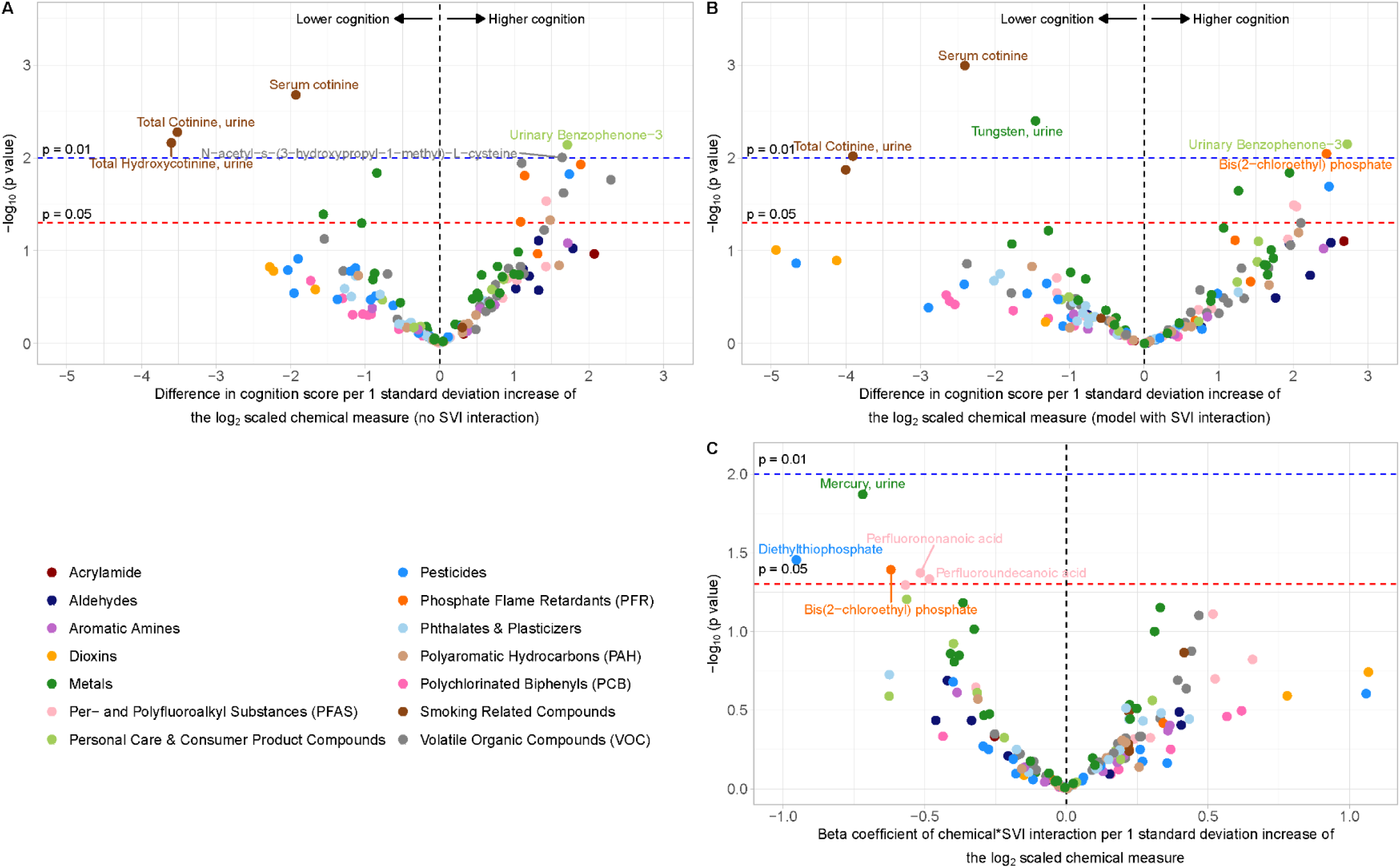
Volcano plots depicting (A) marginal associations between chemical exposures and Digit Symbol Substitution Test (DSST) scores in models without SVI interaction terms; (B) associations between chemical exposures and DSST scores in models with SVI interaction terms; and (C) coefficients of chemical exposure*SVI interactions. The x-axes depict the corresponding model coefficients. The y-axes depict p-values on a -log_10_ scale.

**Table 3.** Summary of survey-weighted linear regression models evaluating the association between chemical exposure biomarkers and Digit Symbol Substitution Test (DSST) scores, with and without Social Vulnerability Index (SVI) interaction, adults aged 60+ in NHANES 1999-2000 and 2011-2014 (overall N = 4,982).

|  |  | Model 1: no interaction |  |  | Model 2: with interaction |  |  |  |  |  |
| --- | --- | --- | --- | --- | --- | --- | --- | --- | --- | --- |
|  |  | Chemical marginal effect estimate |  |  | Chemical effect estimate (SVI = 0) |  |  | Chemical:SVI interaction |  |  |
| Factor | n | β (95% CI) | p-value (unadj) | p-value (adj) | β (95% CI) | p-value (unadj) | p-value (adj) | β (95% CI) | p-value (unadj) | p-value (adj) |
| Metals |  |  |  |  |  |  |  |  |  |  |
| Mercury, blood (ug/L) | 2439 | 0.56 (-0.23, 1.35) | 0.18 | 0.57 | 1.26 (0.27, 2.26) | 0.02 | 0.66 | -0.36 (-0.73, 0.00) | 0.07 | 0.75 |
| Cadmium, urine (ng/mL) | 1549 | -1.56 (-2.99, -0.13) | 0.04 | 0.43 | -1.77 (-3.72, 0.17) | 0.08 | 0.75 | 0.10 (-0.42, 0.62) | 0.71 | 0.95 |
| Mercury, urine (ug/L) | 1093 | 0.50 (-0.40, 1.40) | 0.29 | 0.67 | 1.95 (0.56, 3.34) | 0.01 | 0.53 | -0.72 (-1.21, -0.22) | 0.01 | 0.53 |
| Tungsten, urine (ng/mL) | 1573 | -0.84 (-1.48, -0.21) | 0.01 | 0.23 | -1.46 (-2.37, -0.54) | 0.004 | 0.53 | 0.33 (-0.01, 0.68) | 0.07 | 0.75 |
| Per- and Polyfluoroalkyl Substances (PFAS) |  |  |  |  |  |  |  |  |  |  |
| Perfluorononanoic acid (ng/mL) | 1443 | 1.03 (-0.54, 2.61) | 0.21 | 0.57 | 2.04 (0.24, 3.84) | 0.03 | 0.75 | -0.52 (-0.99, -0.04) | 0.04 | 0.75 |
| Perfluorooctanoic acid (ng/mL) | 1384 | 1.43 (0.20, 2.65) | 0.03 | 0.33 | 0.90 (-1.25, 3.06) | 0.42 | 0.94 | 0.24 (-0.42, 0.90) | 0.48 | 0.94 |
| Perfluoroundecanoic acid (ng/mL) | 1443 | 0.91 (-0.44, 2.26) | 0.20 | 0.57 | 2.01 (0.27, 3.75) | 0.03 | 0.75 | -0.48 (-0.94, -0.03) | 0.046 | 0.75 |
| Personal Care & Consumer Product Compounds |  |  |  |  |  |  |  |  |  |  |
| Benzophenone-3, urine (ng/mL) | 1134 | 1.71 (0.61, 2.81) | 0.007 | 0.23 | 2.72 (1.04, 4.41) | 0.007 | 0.53 | -0.56 (-1.10, -0.03) | 0.06 | 0.75 |
| Pesticides |  |  |  |  |  |  |  |  |  |  |
| 3-phenoxybenzoic acid, urine (ug/L) | 1047 | 1.74 (0.47, 3.00) | 0.01 | 0.23 | 2.48 (0.58, 4.38) | 0.02 | 0.66 | -0.40 (-1.00, 0.20) | 0.21 | 0.90 |
| Diethylthiophosphate, urine (ug/L) | 487 | -0.62 (-1.92, 0.68) | 0.39 | 0.71 | 0.98 (-0.61, 2.58) | 0.29 | 0.90 | -0.95 (-1.56, -0.35) | 0.04 | 0.75 |
| Phosphate Flame Retardants |  |  |  |  |  |  |  |  |  |  |
| Bis(1,3-dichloro-2-propyl) phosphate, urine (ug/L) | 957 | 1.14 (0.30, 1.98) | 0.02 | 0.23 | 1.22 (-0.05, 2.49) | 0.08 | 0.75 | -0.04 (-0.61, 0.52) | 0.88 | 0.99 |
| Bis(2-chloroethyl) phosphate, urine (ug/L) | 963 | 1.08 (0.08, 2.08) | **0.049** | 0.44 | 2.44 (0.81, 4.08) | **0.01** | 0.53 | -0.62 (-1.16, -0.08) | **0.04** | 0.75 |
| Diphenyl phosphate, urine (ug/L) | 967 | 1.89 (0.57, 3.21) | **0.01** | 0.23 | 1.43 (-0.75, 3.60) | 0.21 | 0.90 | 0.22 (-0.51, 0.95) | 0.56 | 0.94 |
| **Polyaromatic Hydrocarbons (PAH)** |  |  |  |  |  |  |  |  |  |  |
| 1-phenanthrene (ng/L) | 1082 | 1.48 (0.12, 2.84) | **0.047** | 0.44 | 2.07 (0.01, 4.13) | 0.06 | 0.75 | -0.31 (-0.85, 0.22) | 0.27 | 0.90 |
| **Smoking Related Compounds** |  |  |  |  |  |  |  |  |  |  |
| Cotinine, serum (ng/mL) | 4687 | -1.93 (-3.06, -0.80) | **0.002** | 0.23 | -2.40 (-3.70, -1.10) | **0.001** | 0.30 | 0.22 (-0.21, 0.65) | 0.32 | 0.90 |
| Total Cotinine, urine (ng/mL) | 535 | -3.52 (-4.89, -2.15) | **0.005** | 0.23 | -3.91 (-5.40, -2.41) | **0.01** | 0.53 | 0.22 (-0.38, 0.82) | 0.52 | 0.94 |
| Total Hydroxycotinine, urine (ng/mL) | 535 | -3.60 (-5.08, -2.12) | **0.007** | 0.23 | -4.00 (-5.71, -2.30) | **0.01** | 0.53 | 0.22 (-0.48, 0.93) | 0.58 | 0.94 |
| **Volatile Organic Compounds (VOC)** |  |  |  |  |  |  |  |  |  |  |
| 2-Methylhippuric acid (ng/mL) | 1022 | 1.66 (0.34, 2.98) | **0.02** | 0.29 | 1.96 (-0.16, 4.09) | 0.09 | 0.75 | -0.17 (-0.81, 0.47) | 0.61 | 0.94 |
| m-/p-Xylene, blood (ng/ml) | 1442 | 1.10 (0.33, 1.87) | **0.01** | 0.23 | 0.75 (-0.53, 2.02) | 0.26 | 0.90 | 0.26 (-0.42, 0.94) | 0.46 | 0.94 |
| N-acetyl-S-(2-cyanoethyl)-L-cysteine, urine (ng/mL) | 1061 | 2.29 (0.58, 4.01) | **0.02** | 0.23 | 2.10 (0.15, 4.05) | 0.05 | 0.75 | 0.09 (-0.48, 0.66) | 0.76 | 0.96 |
| N-acetyl-s-(3-hydroxypropyl-1-methyl)-L-cysteine, urine (ng/mL) | 1068 | 1.64 (0.52, 2.76) | **0.01** | 0.23 | 1.31 (-0.41, 3.02) | 0.15 | 0.82 | 0.17 (-0.43, 0.77) | 0.59 | 0.94 |

Model 1: DSST ~ chemical + SVI + age + sex + smoking status + blood cotinine + urinary creatinine + seafood consumption + cycle
Model 2: DSST ~ chemical + SVI + chemical\*SVI + age + sex + smoking status + blood cotinine + urinary creatinine + seafood consumption + cycle
Chemical exposure, blood cotinine, and urinary creatinine were log<sub>2</sub> transformed and z-score standardized
P-values adjusted by controlling for the False Discovery Rate (FDR)

### 3.3. Interaction Models

When an interaction term between SVI and each chemical biomarker was included in the regression models, the main effects of chemicals (when SVI=0) were relatively consistent (**Figure 2, Panel B; Table 3**). In addition, four new biomarkers that were not identified in the marginal association models showed significant negative interactions with SVI in relation to cognitive function: mercury (urine), diethylthiophosphate (urine), perfluorononanoic acid (PFNA) (serum), and perfluoroundecanoic acid (PFUnA) (serum) (**Figure 2, Panel C; Table 3**). The main effect terms for these four chemicals were positive (**Table 3**), suggesting that adverse effects on cognitive function may occur only among individuals experiencing social vulnerability. For example, among individuals with high SVI (median of upper tertile, 4.5), a doubling of diethylthiophosphate (urine) was associated with ‒3.99 points difference in DSST score (95% CI: ‒6.68, ‒1.30), while the association was not statically significant among individuals with low SVI (median of lower tertile, 1) (β=0.62, 95% CI: ‒1.04, 2.28) (**Figure 3; Supplemental Table 6**). Full results of the interaction models are in **Supplemental Table 7**.

**Figure 3.**
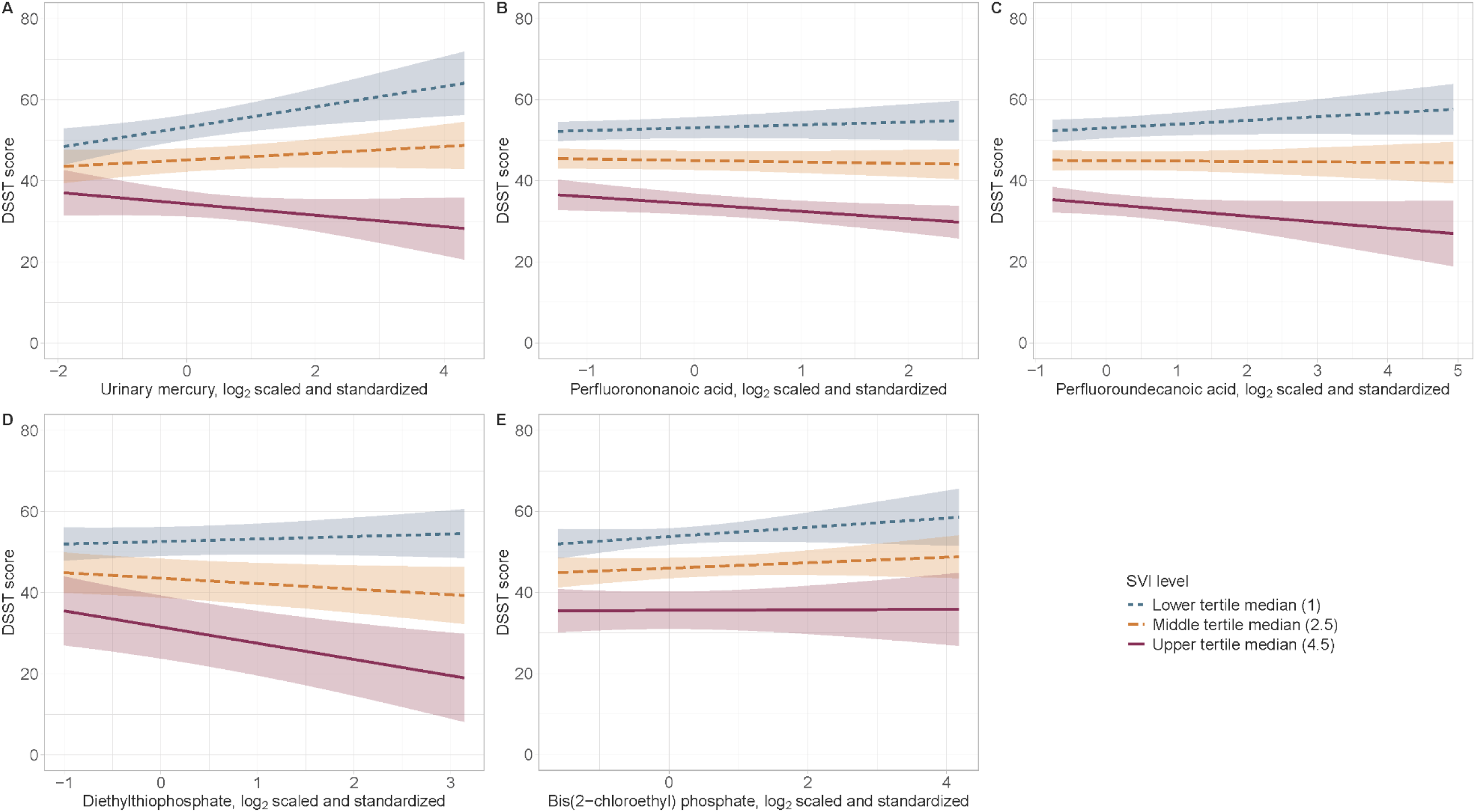
Interaction plots depicting the associations between chemical exposures and Digit Symbol Substitution Test (DSST) scores, fixed at median values of each tertile of the Social Vulnerability Index (SVI). The five chemical exposures featured have significant (unadjusted p<0.05) interaction with SVI, shown in Figure 2 panel C.

### 3.4. Sensitivity Analysis

When restricting the analysis to those with normal kidney function (overall N=3,593), diethylthiophosphate, PFNA, and PFUnA remained significantly associated with DSST score in interactions with SVI (**Supplemental Figure 2; Supplemental Table 8; Supplemental Table 9**). Two additional PFAS, perfluorohexane sulfonic acid (PFHxS) and perfluorodecanoic acid (PFDA), showed significant negative interactions with SVI.

When binary MCI was used as the outcome, four chemicals were identified with *p*<0.01: cotinine (serum) and total cotinine (urine) were associated with higher prevalence of MCI, whereas zinc (serum) and perfluorooctanoic acid (serum) were associated with lower prevalence of MCI in the marginal association models (**Supplemental Figure 3, Panel A; Supplemental Table 10**). In interaction models, the main effects were consistent (**Supplemental Figure 3, Panel B**) and six new chemicals that were not identified in the marginal association models showed significant interactions with SVI. Selected chemicals with *p*<0.01 include cadmium (blood) and bis(1-chloro-2-propyl) phosphate (urine) (**Supplemental Figure 3, Panel C; Supplemental Table 11**).

**Supplemental Figure 4** and **Supplemental Table 12** depict the results of the 2-df Wald test. A total of 25 exposure biomarkers had an unadjusted test p-value <0.05, and the top four (total cotinine, total hydroxycotinine, pentanaldehyde, and serum cotinine) remained significant after FDR adjustment.

## 4. DISCUSSION

To the best of our knowledge, this is the first study to conduct a systematic evaluation of the interactions between large scale environmental chemical biomarkers and a non-genetic susceptibility factor, social vulnerability, on cognition using the exposome-wide interaction study (ExWIS) framework. Using the representative US population data from the NHANES, we identified four additional chemicals, mercury (urine), diethylthiophosphate (urine), PFNA (serum), and PFUnA (serum), that may not have been identified if the analysis did not consider the interaction with SVI. Thus, our framework uncovered the disproportionate effects of these chemicals on the cognitive ability of populations with high social vulnerability. This hypothesis-generating ExWIS approach may be broadly applicable in other cohorts and with other health outcomes.

Social vulnerability is defined as the susceptibility of individuals or social groups to the impacts of external stressors and their ability to cope with them (Cutter et al., 2003). Long-term exposure to social stressors, such as racial and ethnic minority status, low educational attainment, and poverty, may increase the risk for cognitive impairment and other neurodegenerative diseases by promoting chronic stress (Herman et al., 2016; Sapolsky, 1996). A growing body of evidence suggests social stressors may also interact with environmental neurotoxicants (Casey et al., 2023). For example, children with more social stressors characterized by residential segregation and low socioeconomic status had larger negative effects of lead and other metals on neurocognitive outcomes (Bravo et al., 2022; Lucchini et al., 2019; Wylie et al., 2024). Older adults who reported higher levels of psychosocial stress showed worse cognitive score in relation to cumulative lead exposure (Peters et al., 2010). Adversity characterized by socioeconomic difficulties and stressful life events potentiated adverse effects of organophosphate pesticide exposure on child IQ (Stein et al., 2016).

We hypothesize that social vulnerability may modify the neurotoxic effects of environmental chemicals by increasing allostatic load. Allostatic responses are essential physiological processes to adapt to environmental perturbations through activating the hypothalamic-pituitary-adrenal (HPA) axis and stimulating the production of corticosteroids (Karatsoreos and McEwen, 2011). Repeated or prolonged activation or dysregulation of the body’s allostatic responses can lead to a cumulative burden of chronic stress, known as allostatic load (McEwen, 1998; Morello-Frosch and Shenassa, 2006). Allostatic load captures the cumulative ‘wear and tear’ on the body, leading to greater susceptibility to disease and reduced reserve capacity to cope with future stressors (Schwartz et al., 2011). Environmental chemical exposures, such as metals and PFAS, have been associated with allostatic load (Bashir and Obeng-Gyasi, 2022; Halabicky et al., 2023; Souza-Talarico et al., 2017). Additionally, higher allostatic load has been suggested to amplify the adverse effects of lead on blood pressure. A study using the NHANES data found stronger associations between blood lead and systolic and diastolic blood pressure only in the group with high allostatic load but not in the low allostatic load group (Zota et al., 2013).

In the present study, we identified four chemicals–mercury, diethylthiophosphate, PFNA, and PFUnA–that may have potential neurotoxic effects only among individuals with high social vulnerability. Mercury is a well-established neurotoxicant (Branco et al., 2021), and experimental studies have consistently supported its neurotoxic properties (ATSDR, 2024). However, epidemiologic findings have been inconsistent. Early studies on occupational cohorts (such as workers in fluorescent lamp factories and dental professionals) found that exposed workers exhibited poorer cognitive performance compared to unexposed controls (Hilt et al., 2009; Milioni et al., 2017; Sletvold et al., 2012). Studies of the general population exposed to low levels of mercury, however, showed null associations (Lu et al., 2023) or even protective associations (Li et al., 2024; Morris et al., 2016). These discrepancies may be attributed to differences in exposure sources (methylmercury from fish consumption in the general population vs. elemental mercury through occupational exposure) and potential negative confounding effects by beneficial nutrients, such as omega-3 fatty acids, through the shared exposure source by fish consumption (Choi et al., 2008). To account for this, we adjusted for fish consumption as an *a priori* confounder. In fact, we also found non-significant positive marginal effect estimates for blood total mercury and urinary mercury in the marginal association models, as well as significant main effect estimates (when SVI=0) in the interaction models (**Table 3**). Our findings suggest that differential susceptibility to mercury neurotoxicity by social vulnerability may be another potential explanation for the inconsistent epidemiologic findings regarding the association between mercury and neurological outcomes.

Diethylthiophosphate is one of the dialkyl phosphate (DAP) metabolites commonly used as a biomarker for organophophorus pesticides (OPs) exposure (Bravo et al., 2002). OPs are broadly categorized into two main types: dimethyl OPs and diethyl OPs, both of which are widely used to control insects by inhibiting acetylcholinesterase, a cholinergic enzyme responsible for hydrolyzing the neurotransmitter acetylcholine (Trang and Khandhar, 2025). Diethyl OPs, including chlorpyrifos, parathion, malathion and diazinon, are metabolized through oxidation, hydrolysis, and dearylation, and detected as diethylphosphate and diethylthiophosphate (Barr et al., 2004). Beyond acetylcholinesterase inhibition, several additional mechanisms have been proposed to contribute to OP’s neurotoxicity, such as neuroinflammation, oxidative stress and mitochondrial dysfunction, disruptions in cytoskeletal proteins and axonal transport, and alterations in Alzheimer’s disease related pathways (e.g., aberrant amyloid precursor protein processing, beta-amyloid accumulation, tau protein phosphorylation) (Farkhondeh et al., 2020; Guignet and Lein, 2019; Yadav et al., 2024). Robust evidence links OPs exposure and adverse neurobehavioral and neurocognitive effects in occupational settings (Muñoz-Quezada et al., 2016; Ross et al., 2013). However, information on associations between OPs as measured by DAP metabolites and neurocognitive outcomes among non-occupationally exposed adults is limited. Most studies have focused on prenatal or early childhood exposures and neurocognitive outcomes in children. While some studies reported significant associations between urinary DAP concentrations and adverse neurodevelopmental outcomes (Bouchard et al., 2011; Engel et al., 2011; Gunier et al., 2017; Kongtip et al., 2017; Suwannakul et al., 2021), others found no associations (Cartier et al., 2016; Donauer et al., 2016). Recent studies from the Center for the Health Assessment of Mothers and Children of Salinas (CHAMACOS) study found that higher prenatal urinary DAP concentrations were associated with changes in brain imaging outcomes– including regional connectivity (in males only) and activation patterns–during adolescence (Gao et al., 2024; Sagiv et al., 2024). Notably, an earlier CHAMACOS study found that the negative association between prenatal urinary DAP concentrations and child IQ was stronger among children experiencing adversity, such as low income, food insecurity, maternal depression, stressful life events, and family conflict (Stein et al., 2016), which is consistent with the current findings.

PFNA and PFUnA are long-chain perfluoroalkyl carboxylic acids with perfluoroalkyl chains containing nine and eleven carbons, respectively (Buck et al., 2011). Long-chain PFAS, also known as forever chemicals due to their persistence in the environment and in the human body, are known to cross the blood-brain barrier (BBB) and to accumulate in the brain and cerebrospinal fluid (Delcourt et al., 2023; Perez et al., 2013). Experimental studies with perfluorooctane sulfonic acid (PFOS) in mice observed astrocyte damage and increased BBB permeability (Yu et al., 2020). PFAS can also disrupt lipid homeostasis via activation of peroxisome proliferator-activated receptor-alpha (PPARα) and lead to neuroinflammation, mitochondrial dysfunction, protein binding interference, oxidative stress, and alterations in calcium homeostasis and related signaling pathways (Gardener et al., 2025). However, epidemiologic evidence linking PFAS exposure to neurological outcomes is limited and has produced mixed results. While a few studies have reported adverse neurological effects of PFOS (Lefèvre-Arbogast et al., 2024; Park et al., 2021), most other studies have found no association or even positive associations between PFAS and cognitive function (Gallo et al., 2013; Power et al., 2013; Shrestha et al., 2017). In the present study, we also found non-significant, positive marginal associations of PFNA and PFUnA with DSST scores (**Table 3, model 1**). Park et al., (2021) suggested that such positive, protective associations between PFAS and cognitive function may be explained by confounding by fish consumption (a source of PFAS similar to the mercury case above) and effect modification by kidney function. In our analysis, fish consumption was included as a primary confounder. When analyses were restricted to participants with normal kidney function, not only PFNA and PFUnA but also two additional PFAS compounds–perfluorohexane sulfonic acid (PFHxS) and perfluorodecanoic acid (PFDA)–showed significant negative interactions with SVI at *p*<0.05, even though their main effect terms were positive (**Supplemental Figure 3**). These findings suggest that the adverse neurotoxic effects of PFAS may become evident only among individuals experiencing social vulnerability and may not be observed in those with fewer social and economic risk factors. As a hypothesis-generating study, these findings warrant further investigation in well-designed epidemiologic studies.

We conducted a 2-df test as an alternative and sensitivity analysis. This approach compares models with and without two additional terms: the main effect of chemical and the interaction between chemical and SVI. The 2-df test evaluates whether adding these terms significantly improves the model’s goodness-of-fit. This joint test is widely used in genome-wide association studies to increase the power in gene discovery, particularly when both main and interaction effects may contribute to the outcome (Kraft et al., 2007). The 2-df test generally has greater statistical power than marginal association tests when the effect of exposure is predominantly present in individuals with a particular effect modifier, but can be less powerful if the interaction term is null (Boonstra et al., 2016). It is important to note that a statistically significant result with the 2-df test may be driven by either a substantial main effect, a substantial interaction effect, or both. Thus, exposures with strong main effects but no interaction can still be detected by this approach. In our analysis, most chemicals identified using the joint 2-df test overlap with the chemicals with significant main effects but not necessarily strong interaction effects (**Table 3**). However, our main objective was to identify chemicals whose effects are primarily seen in susceptible populations (i.e., individuals with high SVI), rather than chemicals that can be detected through standard marginal association tests.

Several limitations should be considered. First, our study may be underpowered due to the relatively small sample size. Testing for statistical interactions generally requires substantially larger samples compared to analyses of main (marginal) associations. Limited power may explain why our results did not achieve statistical significance after correction for multiple comparisons. Instead, results are presented based on conventional significance thresholds, *p*<0.05 and 0.01. Without adequate adjustment for multiple testing, we cannot rule out the possibility that some of the observed marginal associations and interactions represent false positives. However, as an exploratory analysis, these findings may still be useful in future hypothesis generation. Second, we constructed a modified SVI based on individual-level vulnerability factors rather than neighborhood-level factors due to restrictions on access to geographic information. Other established measures of social vulnerability, such as the Area Deprivation Index (ADI) (Kind and Buckingham, 2018) and the CDC/ATSDR SVI (CDC/ATSDR, 2024), incorporate neighborhood-level factors rather than individual factors. Although individual and neighborhood-level socioeconomic status are often correlated, individual-level data alone cannot capture the broader social context, including community resources and social support, which may influence an individual’s vulnerability or resilience to external stressors (Zahnow, 2024). Future research should consider a more comprehensive SVI that integrates both individual- and neighborhood-level factors. Third, although our ExWIS approach considered multiple chemicals as the exposome, each chemical was analyzed independently, which may not reflect the real-world exposure scenario. Chemicals as a mixture can have cumulative, synergistic, or antagonistic effects, resulting in different biological responses (Braun et al., 2016; Joubert et al., 2022). For example, PFAS and mercury may exert synergistic neurotoxic effects (Reardon et al., 2023). Consideration of chemical mixtures and chemical-chemical interactions is of particular importance, and several methodological tools, such as Bayesian kernel machine regression (Bobb et al., 2015), weighted quantile sum regression (Carrico et al., 2015), quantile g-computation (Keil et al., 2020), and environmental risk scores (Park et al., 2017), are increasingly being utilized in environmental health research. The scope of the current study was to provide a research framework to identify environmental chemicals whose effects may be more pronounced in socially vulnerable subpopulations, rather than to address the full complexity of mixture effects and susceptibility factors in chemical risk assessment (Payne-Sturges et al., 2021). Fourth, the findings of the present study should be interpreted with caution, as the study was not designed to formally test hypotheses or establish causal relationships. Limitations include potential issues related to causal inference, such as temporality, reverse causation, measurement error, and residual confounding. These concerns are inherent to the agnostic, hypothesis-generating nature of this study. The observed associations should therefore be investigated further in well-designed epidemiologic studies that can address these causal inference challenges. Additionally, future exposome-based risk assessments should incorporate other key susceptibility factors such as age and sex, to identify chemical risk factors and susceptible subpopulations.

In addition, this study has several strengths. It was conducted in a nationally representative population of older adults with survey weights; thus, our findings are generalizable to the non-institutionalized US population. A wide variety of chemical biomarkers were measured in blood and urine using highly sensitive and standardized methods in NHANES, which enabled us to perform an exploratory screening of over 100 exposures. We used DSST score as a measure of cognitive function, which is widely used and may enable replication testing in other study populations. Importantly, our study builds on the emerging and hypothesis-generating ExWAS framework, which evaluates exposures with parallel models in an efficient discovery process. We extend these approaches to newly assess joint effects of exposures and social vulnerability, identifying chemical exposures which may be particularly relevant in certain social contexts. Our pipeline is publicly available and transparent, ensuring this framework can be implemented in future studies.

In conclusion, this study provides a risk assessment framework to identify hidden environmental factors that affect susceptible populations through the ExWIS approach. Environmental factors may not be detected as potential neurotoxicants using the mean effect-based ExWAS approach, underscoring the importance of including interactions for risk assessment. Incorporating social vulnerability or other measures of the social determinants of health into exposome-based risk assessment can provide a more comprehensive understanding of how social and biological factors intersect to drive health disparities (Perry et al., 2024). By accounting for social vulnerability, we can move beyond average population effects and identify subpopulations at higher risk, leading to more targeted and effective public health interventions.

## Data Statement

All NHANES data are publicly available through the US National Center for Health Statistics. Code to produce these analyses and graphics is available through GitHub (https://github.com/bakulskilab/Cognition_SVI_ExWIS).

## Author Contributions

SKP: conceptualization, methodology, supervision, project administration, funding acquisition, writing-original draft, writing-review & editing; EW: data curation, formal analysis, visualization, writing-original draft, writing-review & editing; LAM: visualization, writing-original draft, writing-review & editing; SF: formal analysis, writing-review & editing; LYMM: data curation, visualization, writing-review & editing; XW: writing-review & editing; BM: methodology, writing-review & editing; BLN: writing-review & editing; KMB: supervision, project administration, funding acquisition, writing-review & editing

