## Supplement for "An Exposome-Wide Interaction Study to Identify Hidden Environmental Factors Affecting Susceptible Populations: Application to Chemical-Social Vulnerability Interactions on Cognitive Function"

Supplemental tables and figures

**Supplemental Table 1.** Summary of analytic variables with missing data imputed using Multiple Imputation by Chained Equations (MICE).

| Variable | N missing | Imputation method |
|---|---|---|
| DSST score | 721 | Predictive mean modeling |
| **Social Vulnerability Index components** | | |
| Poverty income ratio | 583 | Predictive mean modeling |
| Education level | 15 | Proportional odds modeling |
| Health insurance | 46 | Polytomous logistic regression |
| Housing tenure | 52 | Polytomous logistic regression |
| Employment | 2 | Logistic regression |
| Food security | 57 | Proportional odds modeling |
| **Covariates** | | |
| Urinary creatinine (mg/dL) | 139 | Predictive mean modeling |
| Waist circumference (cm) | 370 | Predictive mean modeling |
| Serum cotinine (ng/mL) | 295 | Predictive mean modeling |
| Smoking status | 7 | Polytomous logistic regression |
| Seafood consumption | 444 | Predictive mean modeling |
| Alcohol consumption | 280 | Predictive mean modeling |

**Supplemental Figure 1.** Inclusion and exclusion flowcharts for (A) the chemical exposure biomarkers and (B) the analytic sample in the National Health and Nutrition Examination Survey (NHANES), years 1999-2000, 2011-2012, and 2013-2014.

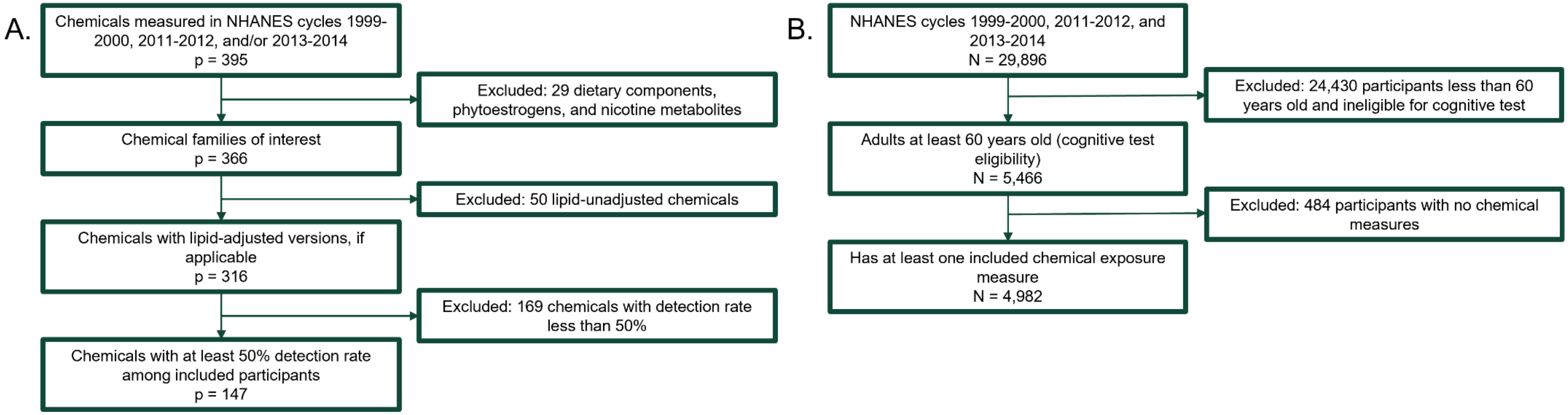

**Supplemental Table 2.** Weighted descriptive statistics of excluded vs. included participants aged 60 years and older, National Health and Nutrition Examination Survey (NHANES) 1999-2000, 2011-2012, and 2013-2014 (N = 5,466). Participants were excluded if they had no chemical measures.

| Variable | Overall (N = 5,466)[1] | Excluded (N = 484)[1] | Included (N = 4,982)[1] | p-value[2] |
|---|---|---|---|---|
| **Digit Symbol Substitution Test (DSST)** | 50.3 (17.4) | 36.6 (14.4) | 50.4 (17.4) | <0.001 |
| Missing | 1,035 | 314 | 721 | |
| **Age (years)** | 69.8 (7.0) | 72.1 (7.8) | 69.8 (7.0) | 0.03 |
| **Sex** | | | | >0.9 |
| Male | 45.0% | 44.5% | 45.0% | |
| Female | 55.0% | 55.5% | 55.0% | |
| **Social Vulnerability Index (SVI)** | 1.9 (1.5) | 2.5 (1.6) | 1.9 (1.5) | 0.02 |
| Missing | 739 | 118 | 621 | |
| **Poverty-income ratio (PIR)** | | | | 0.2 |
| PIR > 1.5 | 73.9% | 63.9% | 74.0% | |
| PIR <= 1.5 | 26.1% | 36.1% | 26.0% | |
| Missing | 696 | 113 | 583 | |
| **Education** | | | | 0.3 |
| Completed high school or above | 76.9% | 71.1% | 77.0% | |
| Did not complete high school | 23.1% | 28.9% | 23.0% | |
| Missing | 24 | 9 | 15 | |
| **Race/ethnicity** | | | | 0.009 |
| Non-Hispanic White | 78.1% | 65.4% | 78.2% | |
| Not Non-Hispanic White | 21.9% | 34.6% | 21.8% | |
| **Health insurance** | | | | 0.1 |
| Private insurance | 61.6% | 49.0% | 61.7% | |
| Government insurance | 33.1% | 45.1% | 33.0% | |
| No insurance | 5.3% | 6.0% | 5.3% | |
| Missing | 73 | 27 | 46 | |
| **Housing tenure** | | | | 0.1 |
| Homeowner | 81.6% | 72.9% | 81.7% | |
| Renter or other | 18.4% | 27.1% | 18.3% | |
| Missing | 81 | 29 | 52 | |
| **Food security** | | | | 0.4 |
| Full security | 88.2% | 90.6% | 88.2% | |
| Less than full security | 11.8% | 9.4% | 11.8% | |
| Missing | 87 | 30 | 57 | |
| **Employment** | | | | 0.006 |
| Employed | 27.0% | 8.6% | 27.3% | |
| Unemployed or retired | 73.0% | 91.4% | 72.7% | |
| Missing | 7 | 5 | 2 | |
| **Serum cotinine (ng/mL)** | 35.9 (105.0) | NA | 35.9 (105.0) | NA |
| Missing | 779 | 484 | 295 | |
| **Smoking status** | | | | 0.5 |

| Variable | Overall (N = 5,466)[1] | Excluded (N = 484)[1] | Included (N = 4,982)[1] | p-value[2] |
|---|---|---|---|---|
| Never smoker | 49.2% | 55.9% | 49.1% | |
| Former smoker | 39.1% | 34.9% | 39.2% | |
| Current smoker | 11.7% | 9.1% | 11.8% | |
| Missing | 14 | 7 | 7 | |
| **Urinary creatinine (mg/dL)** | 104.8 (68.0) | 102.4 (74.4) | 104.8 (67.9) | 0.9 |
| Missing | 578 | 439 | 139 | |
| **Fish and seafood consumption in past 30 days** | | | | 0.3 |
| 0 times | 14.5% | 24.2% | 14.4% | |
| 1-3 times | 33.7% | 37.8% | 33.7% | |
| 4+ times | 51.8% | 38.0% | 51.9% | |
| Missing | 866 | 422 | 444 | |
| **Estimated glomerular filtration rate (eGFR; ml/min/1.73 $m^2$)** | | | | NA |
| eGFR < 60 | 21.3% | NA | 21.3% | |
| eGFR >= 60 | 78.7% | NA | 78.7% | |
| Missing | 772 | 484 | 288 | |
| **NHANES cycle** | | | | <0.001 |
| 1999-2000 | 25.9% | 46.0% | 25.6% | |
| 2011-2012 | 35.9% | 43.6% | 35.8% | |
| 2013-2014 | 38.2% | 10.4% | 38.6% | |

[1]Mean (SD); %; unweighted N missing

[2]Design-based t-test; Pearson's $X^2$: Rao & Scott adjustment

**Supplemental Table 3.** Weighted descriptive statistics of participants aged 60 years and older in National Health and Nutrition Examination Survey (NHANES) years 1999-2000, 2011-2012, and 2013-2014, stratified by cognitive status determined via Digit Symbol Substitution Test (DSST) score (N = 4,982).

| Variable | Mild cognitive impairment (N = 1,493)[1] | No cognitive impairment (N = 2,768)[1] | Missing DSST score (N = 721)[1] | p-value[2] |
|---|---|---|---|---|
| **Digit Symbol Substitution Test (DSST)** | 25.9 (8.3) | 57.1 (12.5) | NA | <0.001 |
| Missing | 0 | 0 | 721 | |
| **Age (years)** | 72.7 (7.0) | 68.5 (6.5) | 73.1 (7.4) | <0.001 |
| **Sex** | | | | 0.4 |
| Male | 46.9% | 44.8% | 42.5% | |
| Female | 53.1% | 55.2% | 57.5% | |
| **Social Vulnerability Index (SVI)** | 3.1 (1.6) | 1.5 (1.3) | 3.0 (1.7) | <0.001 |
| Missing | 191 | 285 | 145 | |
| **Poverty-income ratio (PIR)** | | | | <0.001 |
| PIR > 1.5 | 50.6% | 83.0% | 52.3% | |
| PIR <= 1.5 | 49.4% | 17.0% | 47.7% | |
| Missing | 173 | 276 | 134 | |
| **Education** | | | | <0.001 |
| Completed high school or above | 48.5% | 88.4% | 50.6% | |
| Did not complete high school | 51.5% | 11.6% | 49.4% | |
| Missing | 5 | 1 | 9 | |
| **Race/ethnicity** | | | | <0.001 |
| Non-Hispanic White | 60.3% | 85.7% | 59.6% | |
| Not Non-Hispanic White | 39.7% | 14.3% | 40.4% | |
| **Health insurance** | | | | <0.001 |
| Private insurance | 45.3% | 69.1% | 39.8% | |
| Government insurance | 47.5% | 26.2% | 54.2% | |
| No insurance | 7.2% | 4.7% | 6.0% | |
| Missing | 12 | 15 | 19 | |
| **Housing tenure** | | | | <0.001 |
| Homeowner | 68.2% | 86.7% | 71.9% | |
| Renter or other | 31.8% | 13.3% | 28.1% | |
| Missing | 15 | 23 | 14 | |
| **Food security** | | | | <0.001 |
| Full security | 77.7% | 91.9% | 81.9% | |
| Less than full security | 22.3% | 8.1% | 18.1% | |
| Missing | 22 | 20 | 15 | |
| **Employment** | | | | <0.001 |
| Employed | 13.6% | 33.2% | 11.3% | |
| Unemployed or retired | 86.4% | 66.8% | 88.7% | |
| Missing | 2 | 0 | 0 | |

| Variable | Mild cognitive impairment (N = 1,493)[1] | No cognitive impairment (N = 2,768)[1] | Missing DSST score (N = 721)[1] | p-value[2] |
|---|---|---|---|---|
| **Serum cotinine (ng/mL)** | 54.8 (136.8) | 28.5 (87.8) | 54.3 (138.7) | <0.001 |
| Missing | 93 | 128 | 74 | |
| **Smoking status** | | | | 0.05 |
| Never smoker | 46.7% | 49.1% | 53.1% | |
| Former smoker | 39.2% | 40.0% | 32.9% | |
| Current smoker | 14.1% | 10.8% | 14.0% | |
| Missing | 1 | 3 | 3 | |
| **Urinary creatinine (mg/dL)** | 112.3 (72.5) | 102.9 (66.9) | 103.7 (65.1) | 0.03 |
| Missing | 40 | 24 | 75 | |
| **Fish and seafood consumption in past 30 days** | | | | <0.001 |
| 0 times | 23.7% | 11.6% | 17.9% | |
| 1-3 times | 40.4% | 31.1% | 41.2% | |
| 4+ times | 35.9% | 57.3% | 40.9% | |
| Missing | 129 | 148 | 167 | |
| **Estimated glomerular filtration rate (eGFR; ml/min/1.73 $m^2$)** | | | | <0.001 |
| eGFR < 60 | 31.5% | 17.3% | 31.3% | |
| eGFR >= 60 | 68.5% | 82.7% | 68.7% | |
| Missing | 88 | 127 | 73 | |
| **NHANES cycle** | | | | <0.001 |
| 1999-2000 | 35.2% | 21.8% | 34.3% | |
| 2011-2012 | 30.2% | 37.2% | 36.9% | |
| 2013-2014 | 34.6% | 41.0% | 28.8% | |

[1]Mean (SD); %; unweighted N

[2]Design-based Kruskal-Wallis test; Pearson's $X^2$: Rao & Scott adjustment

Missing DSST scores (N=721) imputed prior to regression models

**Supplemental Table 4.** Chemical descriptive statistics. Attached as separate spreadsheet.

**Supplemental Table 5.** Main analysis results output (no interaction), regardless of significance. Attached as separate spreadsheet

**Supplemental Table 6.** Associations between chemical exposures and Digit Symbol Substitution Test (DSST) scores, fixed at median values of each tertile of the Social Vulnerability Index (SVI) within the subset of participants with that measurement. The five chemical exposures featured have significant (unadjusted p<0.05) interaction with SVI, shown in Figure 2 panel C.

| Factor | Fixed SVI value | β (95% CI) | p-value (unadj) |
|---|---|---|---|
| Mercury, urine | 1 | 2.51 (0.84, 4.17) | 0.005 |
| | 2.5 | 0.83 (-0.45, 2.11) | 0.19 |
| | 4.5 | -1.41 (-3.30, 0.49) | 0.14 |
| Perfluorononanoic acid, urine | 1 | 0.70 (-0.57, 1.97) | 0.27 |
| | 2 | -0.01 (-1.05, 1.02) | 0.98 |
| | 4.5 | -1.80 (-3.39, -0.21) | 0.03 |
| Perfluoroundecanoic acid, urine | 1 | 0.93 (-0.26, 2.11) | 0.12 |
| | 2 | 0.24 (-0.72, 1.21) | 0.61 |
| | 4.5 | -1.47 (-3.12, 0.19) | 0.08 |
| Diethylthiophosphate, urine | 1 | 0.62 (-1.04, 2.28) | 0.41 |
| | 2.5 | -1.36 (-2.91, 0.20) | 0.08 |
| | 4.5 | -3.99 (-6.68, -1.30) | 0.009 |
| Bis(2-chloroethyl) phosphate, urine | 1 | 1.14 (-0.55, 2.82) | 0.18 |
| | 2.5 | 0.67 (-0.63, 1.97) | 0.29 |
| | 4.5 | 0.06 (-1.76, 1.87) | 0.95 |

**Supplemental Table 7.** Main analysis results output (with interaction), regardless of significance. Attached as separate spreadsheet

**Supplemental Figure 2.** Sensitivity analyses with all models restricted to participants with normal kidney function, determined by estimated glomerular filtration rate (eGFR) ≥60 ml/min/1.73 $m^2$. Volcano plots depicting (A) marginal associations between chemical exposures and Digit Symbol Substitution Test (DSST) scores in models without SVI interaction terms; (B) associations between chemical exposures and Digit Symbol Substitution Test (DSST) scores in models with SVI interaction terms; and (C) coefficients of chemical exposure*SVI interactions. The x-axes depict the corresponding model coefficients. The y-axes depict p-values on a $-\log_{10}$ scale.

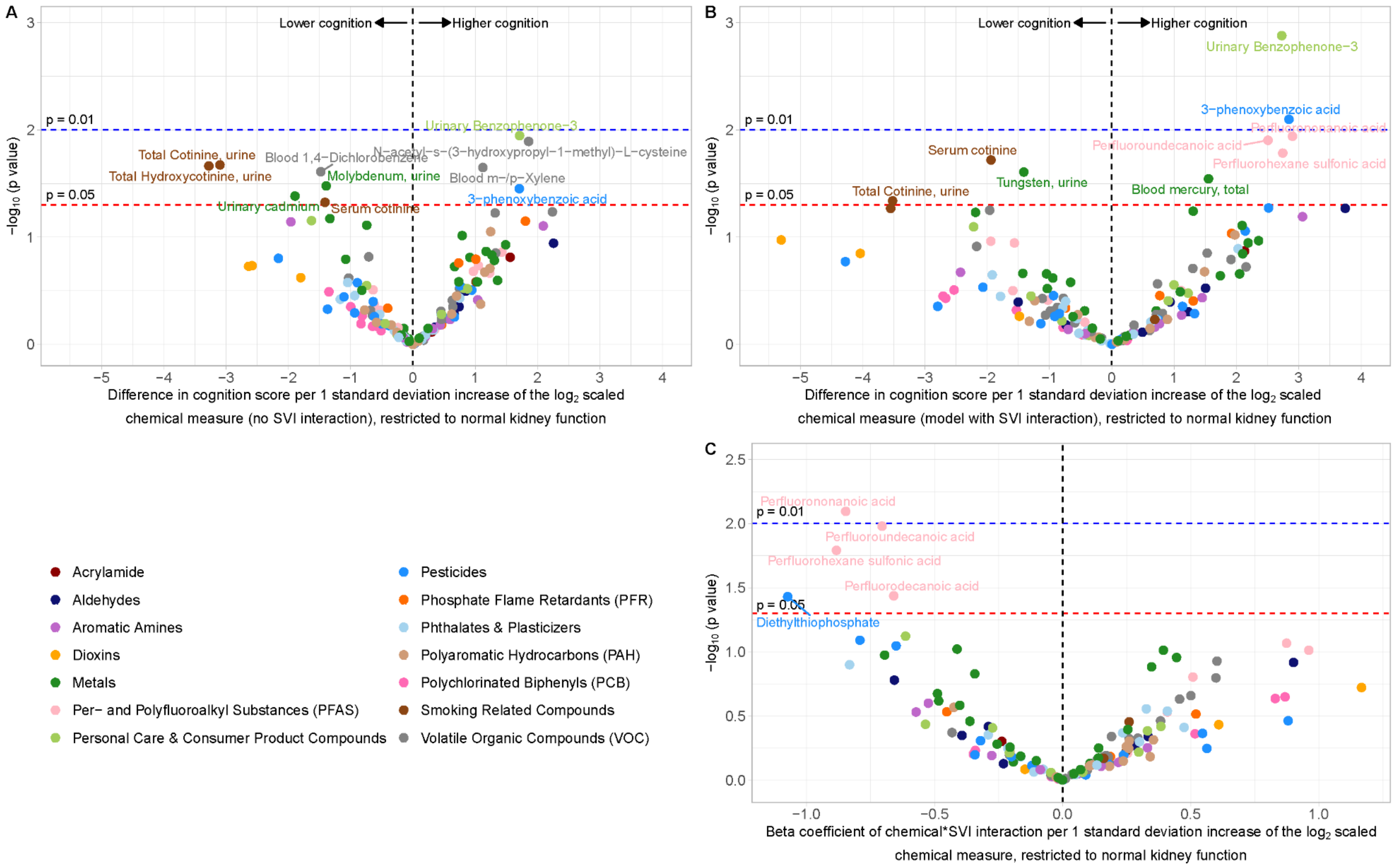


**Supplemental Table 8.** Normal eGFR subset results output (without interaction). Attached as separate spreadsheet

**Supplemental Table 9.** Normal eGFR subset results output (with interaction). Attached as separate spreadsheet

**Supplemental Figure 3.** Sensitivity analyses using binary mild cognitive impairment (MCI) as the outcome. Volcano plots depicting (A) marginal associations between chemical exposures and mild cognitive impairment (MCI) in models without SVI interaction terms; (B) associations between chemical exposures and MCI in models with SVI interaction terms; and (C) coefficients of chemical exposure*SVI interactions. The x-axes depict the corresponding model coefficients. The y-axes depict p-values on a $-\log_{10}$ scale.

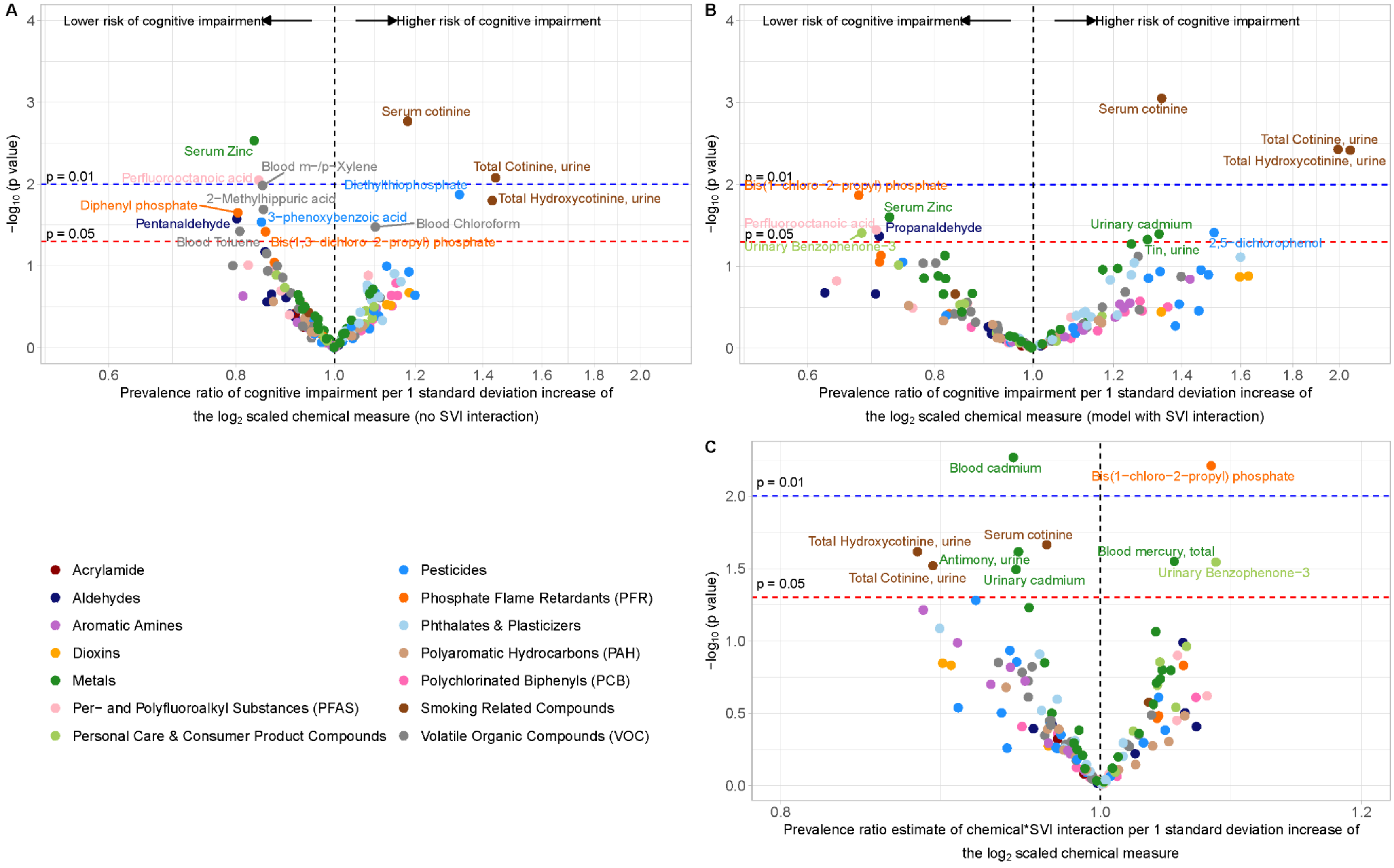


**Supplemental Table 10.** Binary MCI results output (without interaction). Attached as separate spreadsheet

**Supplemental Table 11.** Binary MCI results output (with interaction). Attached as separate spreadsheet

**Supplemental Figure 4.** Manhattan plot of Wald test p-values comparing models without chemical exposure and chemical exposure-SVI interaction terms to models with these terms included. The x-axis depicts chemical exposure family groups. The y-axis depicts p-values on a $-\log_{10}$ scale.

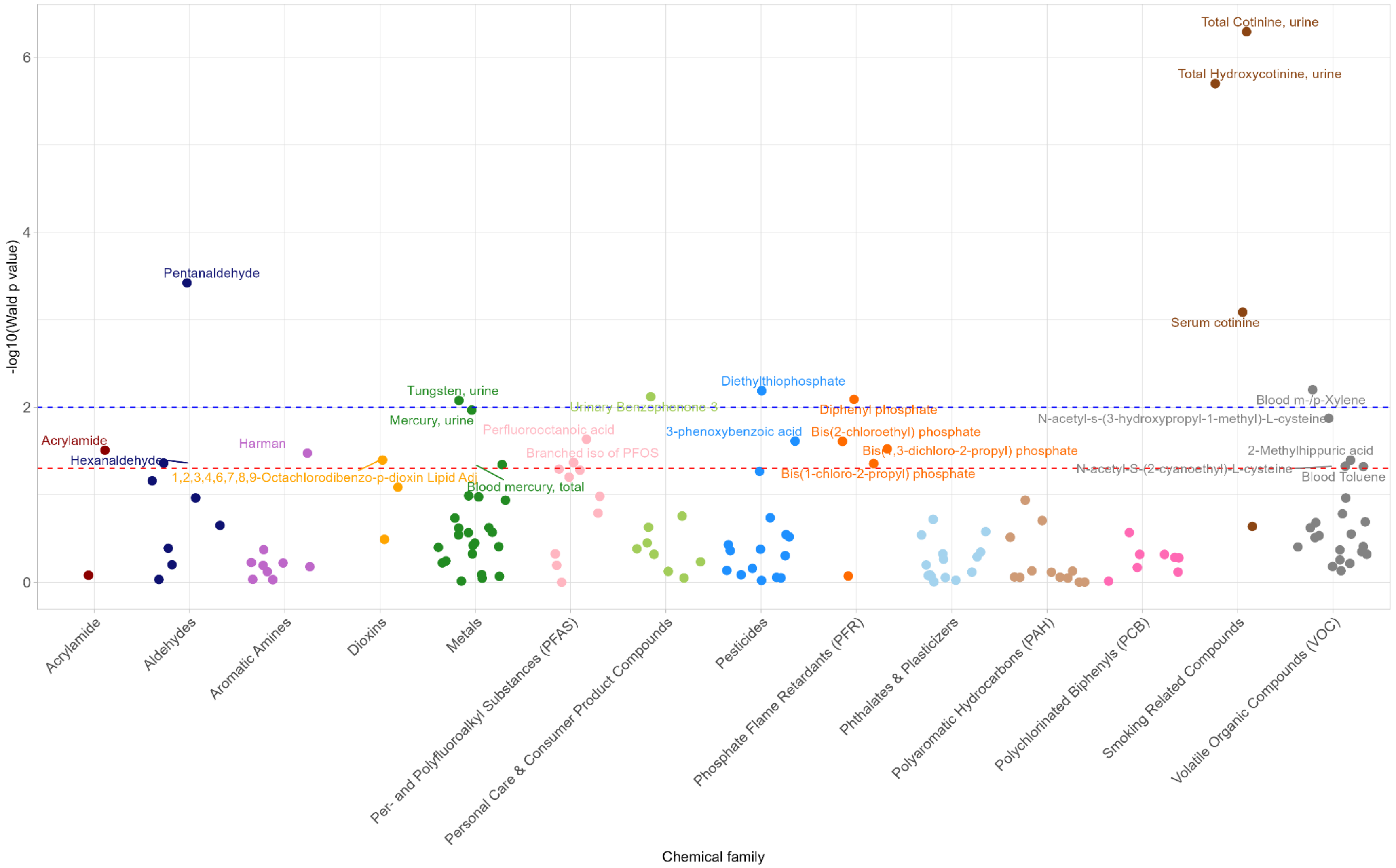


**Supplemental Table 12.** Wald test p-value output, regardless of significance. Attached as separate spreadsheet
